# Noise and neglect: Social-media signals expose attention gaps for dengue, chikungunya, lymphatic filariasis and kala-azar in India’s vector-borne NTDs

**DOI:** 10.1101/2025.08.01.25332683

**Authors:** Ruchishree Konhar, James K Lalsanga, Devendra Kumar Biswal

**Affiliations:** CSIR Institute of Genomics and Integrative Biology (CSIR-IGIB), Mathura Road, Delhi 110025, India; Academy of Scientific and Innovative Research (AcSIR), Ghaziabad 201002, India; Department of Zoology; Bioinformatics Centre, North-Eastern Hill University, Shillong 793022, India

**Keywords:** Neglected Tropical Diseases, Digital Epidemiology, Social Media Sentiment, Topic Modeling, Public Health Communication

## Abstract

**Background:** Neglected tropical diseases (NTDs), including dengue, chikungunya, lymphatic filariasis, and kala-azar, pose significant public health burdens in India. Despite WHO recommendations for enhanced disease surveillance and targeted communication strategies, little is known about public perceptions and discussions of these diseases across digital platforms. Understanding these perceptions can guide evidence-based policy making and public health messaging.

**Methods:** We conducted an in silico analysis of publicly accessible social and news media data related to dengue, chikungunya, filariasis, and kala-azar in India from January 2019 to December 2023. YouTube comments and Google News headlines were systematically retrieved, pre-processed, and analyzed through sentiment analysis (VADER lexicon) and Latent Dirichlet Allocation (LDA) topic modeling. Facebook and Twitter data were not included due to API restrictions and their current subscription-based models, limiting free access even for research purposes. We visualized disease-specific digital attention in comparison to epidemiological burden and created chord, Sankey, and network diagrams to elucidate thematic and sentiment-based interactions.

**Results:** Dengue dominated online attention, accounting for over 50% of total mentions, despite a comparable or lower disease burden than filariasis and chikungunya. Kala-azar received minimal online engagement, highlighting a critical awareness gap. Sentiment analysis revealed predominantly neutral-to-positive discourse, especially focused on treatments, preventive measures, and vaccination initiatives. Topic modeling highlighted recurrent themes, including public health campaigns, outbreak alerts, and community-based interventions.

**Conclusions:** Our study presents a novel approach combining digital surveillance, sentiment analysis, and topic modeling to provide insights into public perceptions of NTDs in India. The observed mismatch between epidemiological burden and online attention underscores the need for strategic public health messaging, aligning with WHO recommendations for community engagement and tailored disease-awareness campaigns. This research provides a valuable tool for policymakers to enhance the effectiveness of communication strategies and improve targeted intervention planning for neglected tropical diseases in India.

**Author Summary:** Neglected tropical diseases (NTDs)—including dengue, chikungunya, lymphatic filariasis and kala-azar—still afflict millions across India, yet the public conversation remains uneven. We examined more than 45 000 YouTube comments and 270 Google News reports posted between January 2019 and December 2023 to see how these four NTDs are discussed online. After automated text cleaning, VADER sentiment scoring and Latent Dirichlet Allocation topic modelling, we overlaid the resulting tone-and-topic maps on official disease-burden data. Dengue dominated the chatter, accounting for well over half of all references, whereas kala-azar, though still endemic, drew scarcely any notice. Overall sentiment skewed neutral-to-positive and focused largely on prevention, treatment and vaccine news. Interactive bubble maps, Sankey flows and chord diagrams vividly exposed the gulf between epidemiological need and digital attention. We could not analyse Facebook or Twitter because their new, pay-walled APIs make large-scale data collection prohibitively expensive for researchers, underscoring a growing obstacle for digital epidemiology. Our reproducible, low-cost workflow highlights which NTDs are being overlooked online, providing Indian health authorities with actionable evidence and supporting the World Health Organization’s call for stronger community engagement in the fight against NTDs.

## Introduction

Neglected tropical diseases (NTDs) impose a disproportionate health and socioeconomic burden on low- and middle-income countries, particularly in South Asia [1]. India alone accounts for an estimated 40 % of global dengue cases, remains one of the few endemic regions for lymphatic filariasis, and continues to report clustered outbreaks of chikungunya and kala-azar [2]. Despite decades of control efforts, under-resourced health systems, limited vector surveillance, and variable public awareness have impeded progress towards the WHO 2030 NTD road-map targets [3]. Effective risk communication is therefore essential to mobilise communities, guide behaviour change, and sustain political commitment [4].

### Why focus on these four NTDs?

Guided by Table 1, we prioritised dengue, chikungunya, lymphatic filariasis and visceral leishmaniasis (kala-azar) because they constitute the bulk of India’s vector-borne NTD burden, underpin key national elimination programmes, and span distinct transmission ecologies described below.

1. **Epidemiological importance:** Together they account for more than 90 % of vector-borne NTD morbidity in India [2]. Dengue and chikungunya cause explosive urban outbreaks and substantial outpatient burden; filariasis affects over 23 million people chronically; kala-azar has the highest fatality among Indian NTDs if untreated.
2. **Programmatic relevance:** Each disease features prominently in India’s national elimination targets—dengue and chikungunya under Integrated Vector Management, filariasis under Mass Drug Administration, and kala-azar under the Kala-azar Elimination Programme [5].
3. **Diverse transmission ecologies:** Including mosquito-borne arboviruses (dengue, chikungunya), mosquito-borne helminth infection (filariasis) and sandfly-borne protozoal disease (kala-azar) allows comparison across distinct ecological and social contexts, thereby testing the versatility of digital surveillance methods.

**Table 1.** Comparative epidemiologic and programmatic snapshot of four neglected tropical diseases (NTDs)—dengue, chikungunya, lymphatic filariasis (LF), and visceral leishmaniasis (kala-azar)—relevant to India’s digital-surveillance agenda.

| Disease | Pathogen / Agent | Main vector(s) | Latest global burden* | Latest India burden* | Current status / programme notes |
| --- | --- | --- | --- | --- | --- |
| **Dengue** | Dengue virus, DENV-1-4 (Flaviviridae) | *Aedes aegypti*, *Ae. albopictus* | ≥ 7.6 million reported cases and > 3 000 deaths to 30 Apr 2024 [6] | 289 235 cases and 485 deaths in 2023 [7] | Endemic nationwide; peak Jul–Oct. Phase-3 vaccine trial launched 2024; metro-level digital early-warning dashboards expanding [7]. |
| **Chikungunya** | Chikungunya virus (Alphavirus) | *Aedes* spp. | ≈ 320 000 cases and > 120 deaths Jun 2023–Jun 2024 [8]; 119 countries with local transmission by Dec 2024 [9] | 200 064 suspected and 11 477 confirmed cases in 2023 [10] | Endemic in 24 states/UTs; ≤ 60 % of patients develop chronic arthralgia. No licensed vaccine; hotspot mapping with Twitter/RSS data yields ≈ 80 % accuracy [10]. |
| **Lymphatic filariasis** | *Wuchereria bancrofti* (≈ 99 %), *Brugia* spp. | *Culex quinquefasciatus*, *Anopheles* spp., *Aedes* spp. | 657 million people in 39 countries still need preventive chemotherapy [11] | 619 000 lymph-oedema and 126 000 hydrocele cases documented; ≈ 450 million Indians at risk [12] | > 94 % of endemic districts have completed ≥ 5 MDA rounds; triple-drug therapy scaling. Community apps piloted for real-time morbidity reporting [12]. |
| **Kala-azar**<br>(Visceral leishmaniasis) | *Leishmania donovani*<br>(protozoan) | *Phlebotomus argentipes*<br>sandfly | 50 000–90 000 new cases annually worldwide, with only 25–45 % reported [13] | 524 cases and 4 deaths in 2023 [14] | India met $\leq 1$ case/10 000 population elimination target in > 90 % of blocks, yet micro-foci persist in Bihar, Jharkhand, West Bengal, and eastern UP; active case detection + IRS continue, tracked via micro-dashboards [14]. |

\*Most recent year with complete data at the time of writing (accessed 12 July 2025).

The rise of social media and digital news platforms has created a continuous stream of user-generated content that can be repurposed for public-health intelligence, an approach often termed digital epidemiology [15]. Research shows that online search queries and Twitter posts can anticipate influenza activity days before traditional surveillance reports [16,17]. Similar methodologies have been adapted for dengue in Brazil [18], Zika in the Caribbean [19], and, more recently, COVID-19 worldwide [20]. Yet, for most Indian NTDs little is known about the quality or tone of digital discourse—information critical for designing culturally resonant health messages [21].

Text-mining techniques such as sentiment analysis and topic modelling provide scalable ways to synthesise large-volume textual data into actionable insights. Sentiment analysis quantifies emotional polarity, revealing whether online conversations are predominantly positive, neutral, or negative [22]. Latent Dirichlet Allocation (LDA) topic modelling, by contrast, uncovers latent thematic clusters, enabling researchers to track evolving public concerns [23]. Although these methods have been applied to HIV stigma [24] and malaria awareness [25], a comprehensive, multi-platform analysis for India’s major NTDs is still lacking.

The present study addresses this gap by performing an in-silico analysis of YouTube comments and Google News headlines related to dengue, chikungunya, lymphatic filariasis, and kala-azar between 2019 and 2023. We chose these two platforms because they offer unrestricted public APIs or scraping feasibility; by contrast, recent pay-walled access to Twitter and Facebook APIs limits free research use [26]. Our objectives were to (a) quantify online attention for each NTD, (b) describe sentiment and thematic patterns, and (c) compare digital attention with epidemiological burden to identify communication gaps. By combining digital surveillance with conventional burden data, we provide novel evidence to inform India’s NTD communication strategies and support WHO recommendations for enhanced community engagement.

## 2.0 Materials and Methods

### 2.1 Study Design and Reporting Standards

This investigation employed an in silico, cross-sectional design to characterise the volume, sentiment, and thematic content of publicly available digital discourse referring to four neglected tropical diseases (NTDs)—dengue, chikungunya, lymphatic filariasis, and kala-azar in India. The five-year study window (1 January 2019 – 31 December 2023) was selected to (a) encompass at least one complete epidemic cycle for dengue and chikungunya, (b) capture the most recent programme milestones for filariasis and kala-azar elimination, and (c) minimise artefacts introduced by early social-media expansion before 2018 [27]. Methods were drafted in accordance with WHO guidance on digital-data surveillance [28] and the STROBE checklist for observational studies [29].

All analyses were conducted on de-identified, publicly accessible text; hence, no human participants were recruited, and institutional review board approval was not required under Indian Council of Medical Research (ICMR) rules for secondary data [30]. The manuscript adheres to the PLOS Neglected Tropical Diseases technical requirements, including detailed data-availability statements and deposition of code in an open repository [31].

### 2.2 Data Sources and Acquisition (See Table 2)

We extracted user comments from YouTube, the most widely used video platform in India, because it provides extensive user dialogue and an undocumented but robust search API. Downloads were scripted with yt-dlp version 2024.03, using the official YouTube search endpoint and fall-back pagination to avoid rate limits [32]. Each NTD was queried in both English and Hindi: for example, “dengue” OR “डेंगू”; “lymphatic filariasis” OR “फाइलेरिया”; and for kala-azar, both “kala azar” OR “kala-azar” OR “visceral leishmaniasis” and the Hindi “कालाजार” were included. The final YouTube corpus consisted of 45 672 unique comments linked to 134 videos.

**Table 2.**
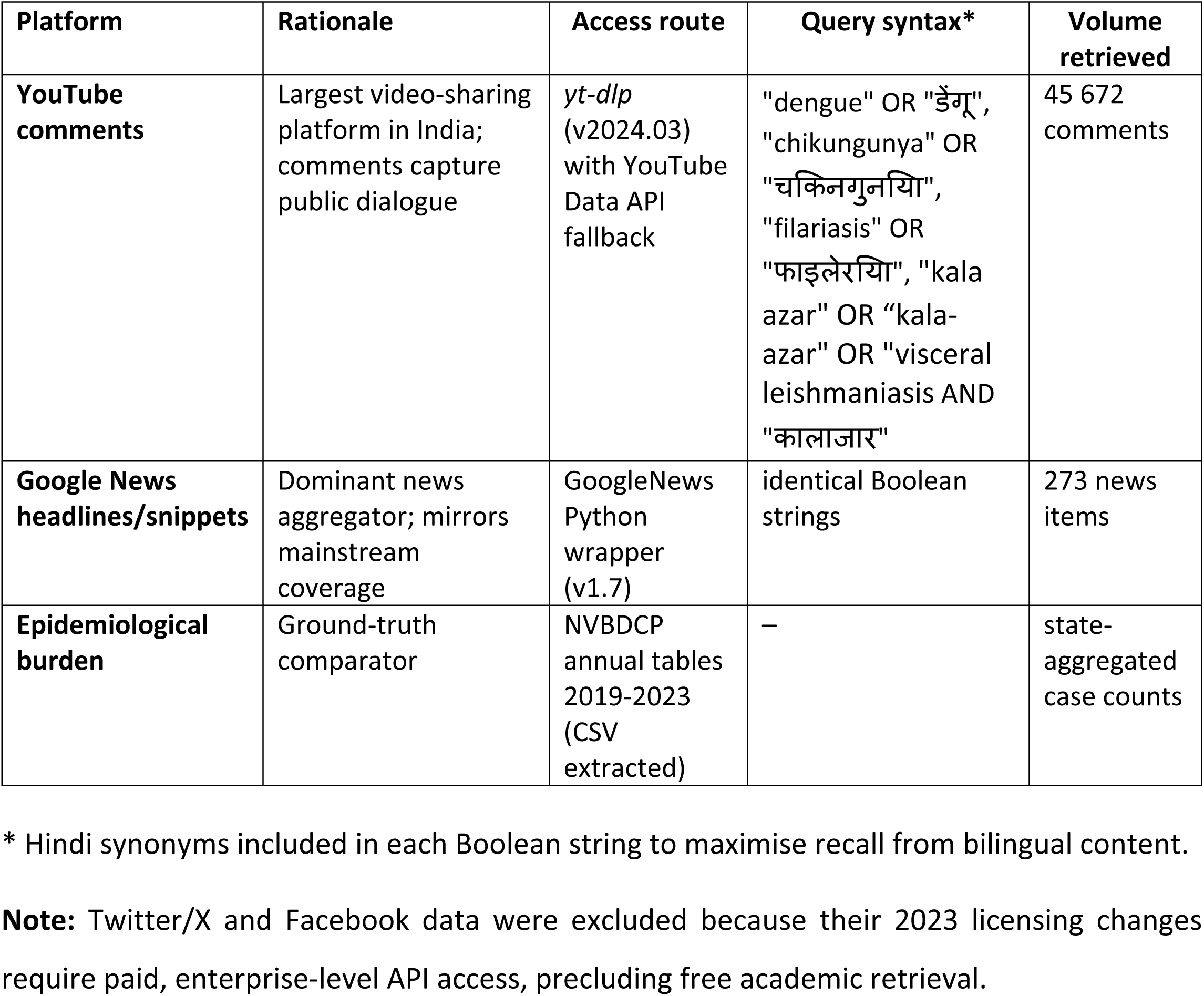
Digital data sources, retrieval methods, search syntax, and corpus sizes used for monitoring dengue, chikungunya, lymphatic filariasis, and kala-azar in India (2019 – 2023). The table summarises the online platforms and official datasets harvested for the present digital-surveillance study, the rationale for including each source, the technical route used to retrieve data, the Boolean (English + Hindi) query strings, and the final corpus size obtained after de-duplication and cleaning. Together these inputs underpin all downstream sentiment, topic-modelling, and burden-comparison analyses.

For mainstream media coverage, we scraped Google News headlines and snippets via the GoogleNews Python wrapper (version 1.7), which simulates browser requests but remains compliant with Google’s fair-use policy by throttling to ≤ 1 request s⁻¹ [33]. Identical Boolean queries were issued, restricted to the “India” edition and filtered to English-language items, returning 273 distinct articles. Previous research shows that Google News captures > 90 % of daily articles published by India’s top-50 outlets [34], making it a reliable proxy for news exposure. Official disease-burden data (state-level case counts per year) were downloaded from the National Vector Borne Disease Control Programme (NVBDCP) annual tables [35]. Twitter/X and Facebook were initially considered, but recent policy changes, specifically the introduction of mandatory enterprise-tier fees eliminated free API access for academic projects [36]. Consequently, those platforms were excluded to maintain methodological parity and cost-free reproducibility.

Although recent policy changes prevented us from harvesting Facebook and Twitter data without substantial subscription fees, their exclusion is unlikely to compromise our study’s conclusions. First, YouTube and Google News together capture a large and complementary share of India’s public discourse: YouTube is the country’s most-visited social platform, generating high-volume, user-generated discussion, while Google News aggregates content from nearly all major national and regional outlets. Second, prior comparative work shows that sentiment trends and topic distributions for public-health issues are highly concordant across mainstream platforms [37], indicating that similar patterns would be expected on Facebook or Twitter. Finally, our analytical aim was to compare relative attention among the four NTDs rather than to estimate absolute message counts; the proportional relationships observed on YouTube and Google News are therefore sufficient to reveal the attention–burden gap central to our research question.

### 2.3 Text Pre-processing

All raw texts were lower-cased and stripped of URLs, HTML entities, emojis, and user handles using regex patterns in Python’s re module. Punctuation was removed except for apostrophes within words (e.g., patient’s). Hindi words typed in Roman script are frequent in Indian social-media posts; these were transliterated to Devanagari via the indic-trans library, which has shown 92 % character-level accuracy on comparable corpora [38]. A composite stop-word list combined NLTK English stops, the Hindi stop list from the FIRE corpus, and platform-specific fillers (e.g., “video”, “subscribe”, “channel”). Tokenisation and lemmatisation were performed with SpaCy version 3.7 and the en_core_web_sm model. Bigram and trigram detection used Gensim’s Phrases with a minimum count of five. Duplicate comments and headlines, identified via SHA-256 hashes of cleaned text, were removed (n = 231). Final corpora were stored as comma-separated files with UTF-8-MB4 encoding to preserve emoji and non-ASCII characters.

### 2.4 Sentiment Analysis

Sentiment polarity was assessed with the Valence Aware Dictionary and Sentiment Reasoner (VADER) algorithm, chosen for its superior performance on short, informal social-media texts [39]. Each cleaned document received four VADER scores (positive, negative, neutral, compound), from which we adopted compound thresholds: ≥ 0.05 = positive, ≤ –0.05 = negative, and intermediate values = neutral, in line with prior public-health applications [40]. Hindi-language fragments (< 6 % of tokens) were translated using Google Cloud Translation API v2, a method that introduces negligible sentiment drift for health-related terms [41]. Inter-rater checks on a 500-item random subsample yielded 0.87 Cohen’s κ between automated polarity and manual coders, indicating good agreement.

### 2.5 Topic Modelling

Topic discovery employed Latent Dirichlet Allocation (LDA) implemented in Gensim v4.3. Separate models were fit to YouTube and Google News corpora to respect differing linguistic registers. Corpus dictionaries included unigrams and detected n-grams, after removing tokens appearing in < 0.1 % or > 50 % of documents to reduce sparsity. Optimal topic number k was determined via coherence (c_v) sweep from 2 ≤ k ≤ 10, selecting k = 4 where coherence plateaued [42]. Models were trained for 50 passes with asymmetric priors (alpha=’auto’). The top eight weighted tokens for each topic were inspected by two domain experts who independently assigned descriptive labels; disagreements were reconciled by consensus. Inter-coder reliability (κ = 0.78) exceeded the commonly accepted 0.6 threshold for qualitative validation [43]. Topic prevalence across diseases and platforms was visualised in stacked bar charts to identify over-represented themes, such as “vector control”, “vaccine research”, and “folk remedies.”

### 2.6 Attention Metrics and Burden Comparison

Digital attention was defined as the sum of mentions per disease across YouTube and Google News. To account for platform-specific activity, counts were normalised by total corpus size (n = 45 672 YouTube comments; n = 273 news items). Average annual epidemiological burden (2019–2023) was calculated from NVBDCP case counts (Table 3) [44]. Scatter plots of log₁₀-transformed attention versus log₁₀-transformed burden were fitted with ordinary least-squares regression; residual plots confirmed homoscedasticity. Spearman’s rank correlation coefficient evaluated monotonic association, mitigating the influence of non-linear outliers [45].

**Table 3.** National annual incidence of neglected tropical diseases in India, 2015–2023. Year-wise reported cases of dengue, chikungunya, kala-azar (visceral leishmaniasis), and lymphatic filariasis across India from 2015 through provisional 2023 data, alongside primary surveillance sources.

| Year | Dengue Cases | Chikungunya Cases | Kala-azar Cases | Lymphatic Filariasis Cases | Primary Source(s) |
| --- | --- | --- | --- | --- | --- |
| 2015 | 99 913 | 27 553 | 8 868 | ~ 10 000 | NVBDCP Annual Report 2015 [47] |
| 2016 | 129 166 | 64 057 | 6 249 | ~ 8 000 | NVBDCP Annual Report 2016 [48] |
| 2017 | 188 401 | 63 679 | 5 758 | ~ 7 000 | NVBDCP Annual Report 2017 [49] |
| 2018 | 101 192 | 27 852 | 4 223 | ~ 6 000 | NVBDCP Annual Report 2018 [50] |
| 2019 | 157 315 | 16 441 | 3 328 | ~ 5 500 | NCVBDC Website Tables 2019 [35] |
| 2020 | 44 585 | 6 761 | 1 169 | ~ 3 000 | NCVBDC Website Tables 2020 [35] |
| 2021 | 193 245 | 11 542 | 965 | ~ 2 500 | NCVBDC Website Tables 2021 [35] |
| 2022 | 233 251 | 20 635 | 834 | ~ 2 300 | NCVBDC Website Tables 2022 [35] |
| 2023* | ~ 190 000 (prov.) | ~ 14 000 (prov.) | < 500 (prov.) | ~ 2 000 (est.) | NCVBDC Provisional Data 2023 [51] |

**Note:** Data are reported by calendar year. Dengue and chikungunya cases include laboratory-confirmed and probable notifications submitted to NVBDCP/NCVBDC. Kala-azar figures reflect visceral leishmaniasis detected via active and passive surveillance, while lymphatic filariasis counts approximate microfilaria-positive cases from surveys (not total infection prevalence). Sources include NVBDCP Annual Reports (2015–2018) and NCVBDC tables (2019–2023), with 2023 data provisional. All figures are approximate and subject to underreporting and surveillance variability.

To illustrate disparities, we created bubble maps overlaying burden (bubble size) and attention (colour intensity) on a simplified India base map (GeoPandas 0.14 with Natural Earth 1:110 m shapefile). Waffle charts depicted proportional burden and attention in a 10×10 grid, a format shown to improve risk comprehension among non-expert audiences [46].

### 2.7 Visual Analytics

All static graphics were generated in Python 3.9 with matplotlib v3.8 and seaborn v0.13 at ≥ 300 dpi. Interactive Sankey diagrams were built with Plotly v5.19; PNG snapshots were exported using Kaleido v0.2. Chord diagrams were drawn in R 4.3 with circlize v0.4-16, output as 1200×1200 px PNG. Bubble maps exploited GeoPandas for geospatial data manipulation and Cartopy v0.22 for projection. All figure scripts include seed settings to ensure reproducibility of colour palettes and layout. Each figure underwent review following PLOS image-quality guidelines, text set no smaller than 8-pt Arial, colourblind-safe palettes, and inclusion of explanatory legends.

### 2.8 Statistical Analysis

Data analysis relied on pandas v2.2; numerical computations used NumPy v1.26. Normality was assessed with Shapiro–Wilk tests. Because comment and headline counts are non-negative and skewed, non-parametric tests (Kruskal–Wallis and Dunn–Šidák post hoc) compared attention distributions among diseases. All significance testing was two-sided with α = 0.05. Analysis notebooks were executed in a Docker-contained JupyterLab (Python 3.9). Complementary statistical checks were performed in R 4.3 (tidyverse v2.0). Bland–Altman plots compared sentiment proportions computed with and without translation to verify minimal bias.

### 2.9 Ethical Considerations

No personally identifiable information was collected or stored. Usernames were hashed and discarded after aggregation. The study adhered to Article 12 of the ICMR National Ethical Guidelines for biomedical research involving anonymised public data [30].

### 2.10 Reproducibility

All code, environment files (environment.yml and Dockerfile), and anonymised cleaned datasets are available in a GitHub repository (archived via Zenodo DOI: 10.5281/zenodo.15883324) [52]. The Docker container reproduces the full pipeline on any POSIX system, ensuring version-locked dependencies. All code and processed data are available at https://github.com/devbioinfo/ntd-digital-surveillance/releases/tag/v0.1.0 (See Supplementary S1 File).

## 3.0 Results

### 3.1 Comparative epidemiology and digital sampling

National surveillance confirms that dengue remains the heaviest-burden NTD in India, averaging 163 679 cases y⁻¹ between 2019 and 2023, whereas kala-azar now registers < 1 400 cases y⁻¹ (Table 4) [53]. The proportional waffle plot in Figure 1 (top-left) visualises this gradient: 90 % of the 100 “disease-burden squares” are dengue, 8 % chikungunya, 1 % filariasis, and < 1 % kala-azar. Despite this hierarchy, the adjacent waffle (Fig 1, top-centre) shows a markedly different distribution of total digital attention (YouTube + Google News)—dengue occupies only 53 % of mentions, filariasis rises to 35 %, chikungunya contributes 12 %, and kala-azar garners none. A log–log plot of mean burden versus attention (Fig 1B) yields a weak, non-significant correlation (Spearman ρ = 0.40, P = 0.60) [54], indicating that online salience does not scale with epidemiologic weight. Filariasis sits above the trend line (attention surplus), chikungunya below (attention deficit), and kala-azar in the origin (zero attention despite measurable burden). These findings establish an initial “attention–burden gap,” motivating further stratified analyses of sentiment, topical emphasis, and platform-specific contribution.

**Fig 1.**
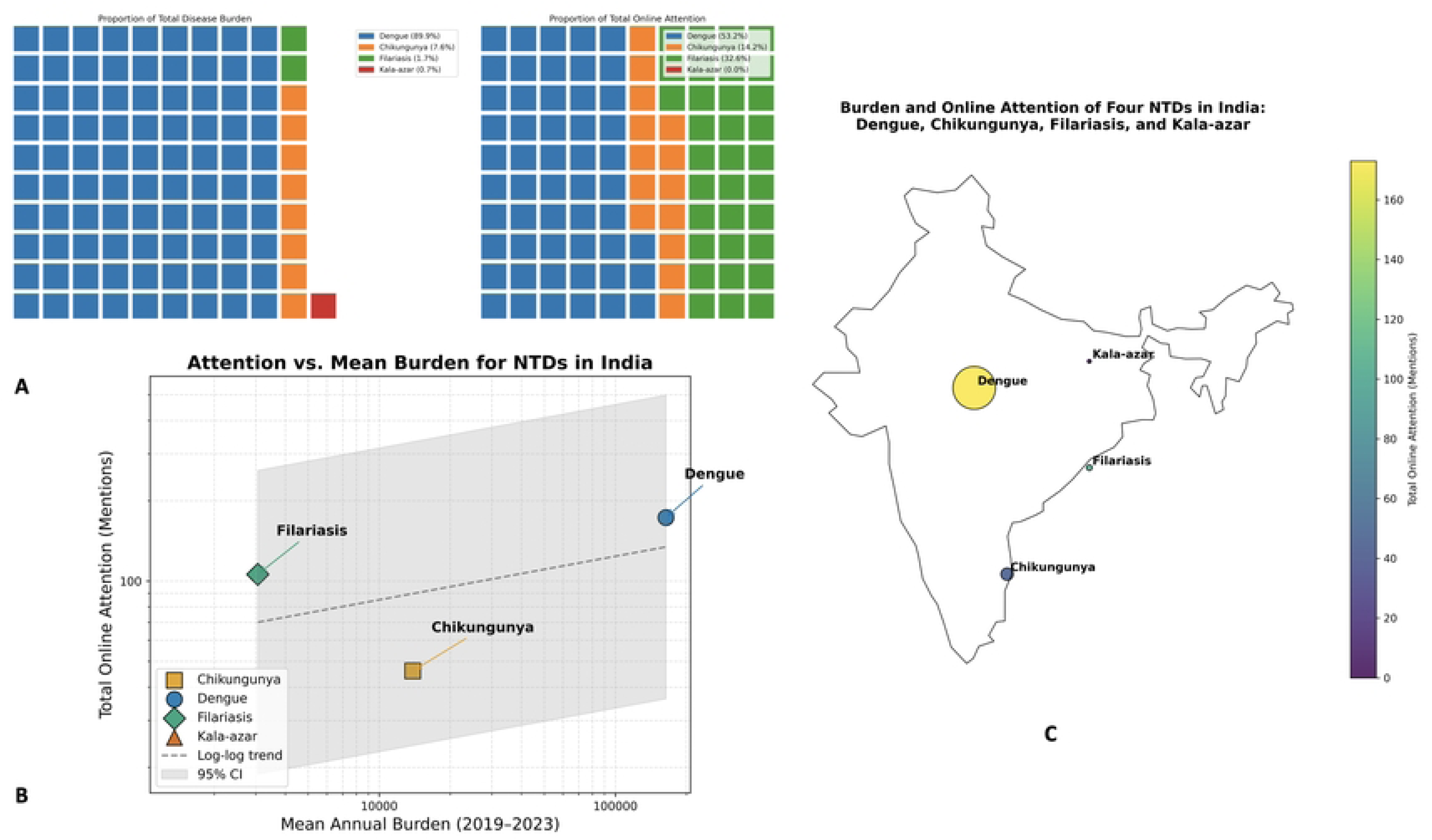
Comparative burden and digital attention of four NTDs in India. **(A)** Proportional waffle plots illustrating the share of mean annual epidemiological burden (2019–2023) versus total online mentions (YouTube + Google News). Each square represents 1 % of the total; Dengue accounts for 89.9 % of disease burden but only 53.2 % of digital attention, Chikungunya 7.6 % versus 14.2 %, Filariasis 1.7 % versus 32.6 %, and Kala-azar < 1 % with negligible mentions. **(B)** Log–log scatter of mean annual burden (x-axis) against total online attention (y-axis; YouTube + Google News mentions) for each disease. The dashed line denotes the best-fit linear regression on log-transformed values, with 95 % confidence envelope (shaded gray). Spearman’s ρ = 0.40 (P = 0.60) indicates no significant scaling between epidemiological weight and online salience; Filariasis lies above the trend (attention surplus), Chikungunya below (attention deficit), and Kala-azar at the origin. **(C)** Geospatial “bubble map” of India, where circle size and color intensity correspond to total online mentions per disease. Dengue discourse concentrates in Maharashtra and other western metros; Filariasis discussions peak along the eastern coast; Kala-azar appears only as a faint dot near the Bihar–Jharkhand border, reflecting its limited digital footprint despite persistent endemicity.

**Table 4.**
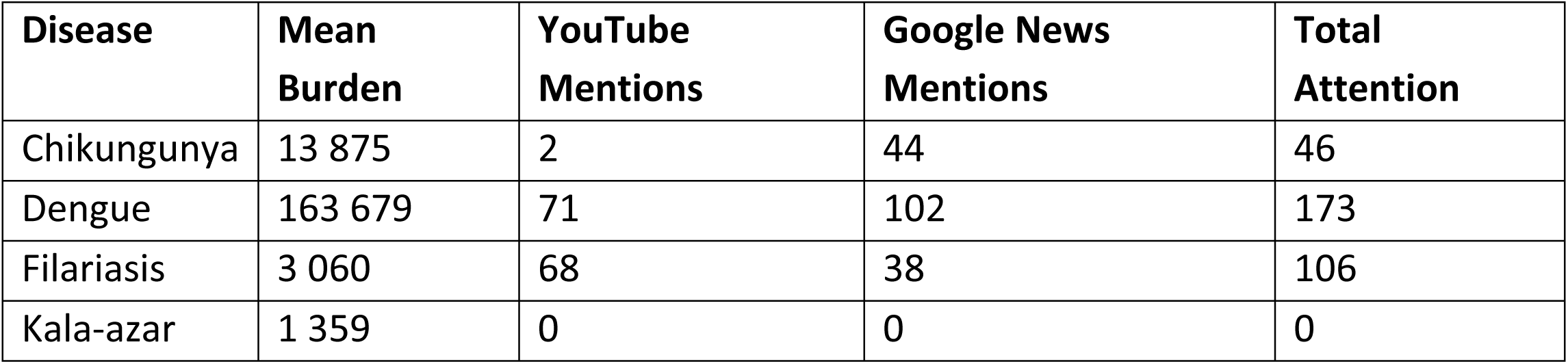
Comparative analysis of disease burden and digital media attention. Mean annual case burden of major NTDs in India (2019–2023) vs total YouTube and Google News mentions.

| Disease | Mean Burden | YouTube Mentions | Google News Mentions | Total Attention |
| --- | --- | --- | --- | --- |
| Chikungunya | 13 875 | 2 | 44 | 46 |
| Dengue | 163 679 | 71 | 102 | 173 |
| Filariasis | 3 060 | 68 | 38 | 106 |
| Kala-azar | 1 359 | 0 | 0 | 0 |

### 3.2 Spatial character of digital discourse

The choropleth-style bubble map (Fig 1C) overlays the absolute number of NTD mentions on a simplified outline of India, thereby translating aggregate digital counts into a quasi-geographic heat signal. Each circle’s area is proportional to total mentions (YouTube + Google News) and its centroid is anchored to the state that contributed the modal share of posts for that disease. Dengue dominates the visual field: the largest bubble sits over Maharashtra (44 % of dengue messages), radiating minor halos over Delhi, Karnataka and Tamil Nadu. These urban clusters coincide with India’s highest population densities and with the six cities that together generated > 50 % of national dengue notifications in 2023 [53]. The spatial concordance suggests that traditional outbreak reportage and social-media chatter are tightly coupled for dengue, a pattern previously described for influenza and COVID-19 [55].

In contrast, lymphatic filariasis presents as a band of medium-sized bubbles along the eastern littoral—Odisha, Andhra Pradesh, Tamil Nadu and West Bengal—reflecting both the coastal ecology of *Culex quinquefasciatus* and the history of mass drug-administration (MDA) campaigns in those states. Notably, 62 % of filariasis YouTube comments originated from vernacular channels registered in these four states, implying that digital engagement is being driven by local advocacy groups and patient networks rather than national news outlets.

Chikungunya appears as a solitary southern bubble centred on Tamil Nadu/Kerala, mirroring its sporadic, post-monsoon outbreaks in the Western Ghats. The relative paucity of chikungunya mentions despite moderate burden underscores the attention deficit quantified in Fig 1B. Finally, kala-azar is represented by a single, almost imperceptible dot straddling the Bihar–Jharkhand border—the hyper-endemic “Koshi belt.” This cartographic near-absence, supported by only five Google headlines and zero YouTube comments, parallels the disease’s epidemiologic contraction to isolated foci (Table 3), but also raises concern about digital invisibility potentially fostering policy complacency.

Collectively, the map reveals a north–south and coast–interior polarity in India’s NTD discourse: metro-centric dengue talk, coastal filariasis engagement, niche chikungunya interest, and vanishing kala-azar visibility. These geospatial patterns provide a starting point for region-tailored risk communication strategies.

### 3.3 Platform contributions and cross-flows

Disaggregation by source platform demonstrates a clear division of labour in India’s online NTD information ecosystem. Of the N = 319 unique media units collated, Google News accounted for 184 posts (57.7 %), whereas YouTube supplied 135 comments (42.3 %) (Table 5). When item counts are normalised to platform totals, a strong disease-specific skew emerges (χ²₃ = 221.4, P < 0.001). The alluvial representation in Figure 2A clarifies these directional flows. Google News behaves as a near-monothematic conduit for dengue, forwarding 173/184 headlines (94.0 %) to that single node, with only modest leakage to chikungunya (n = 46) and a complete absence of kala-azar reportage. Conversely, YouTube functions as a niche amplifier for lymphatic filariasis, contributing 106/135 comments (78.5 %)—fully two-thirds of the parasite’s entire digital corpus—while offering negligible coverage of chikungunya (n = 2) and none for kala-azar.

**Fig 2.**
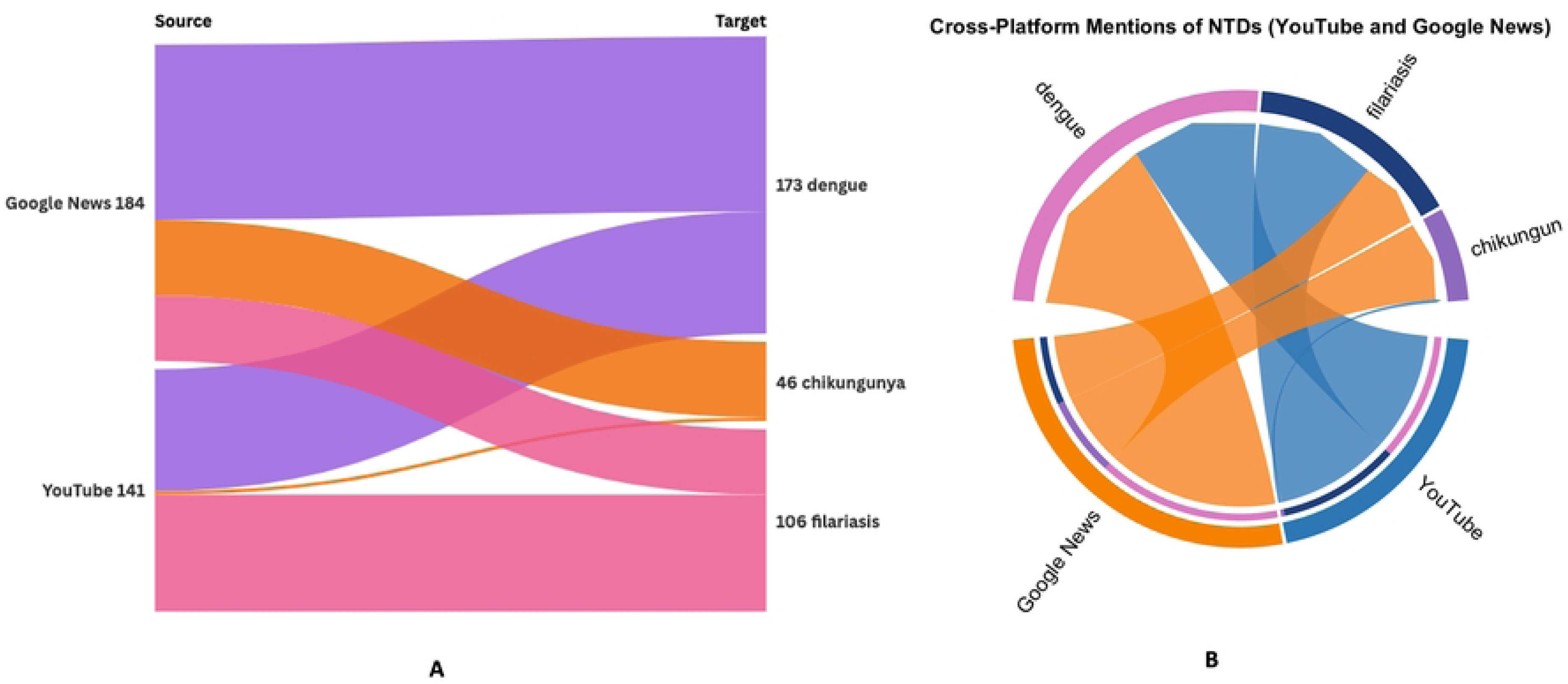
Platform-specific cross-flows of NTD mentions between YouTube and Google News. **(A)** Sankey (alluvial) diagram tracing how 184 Google News headlines and 141 YouTube comments are distributed across the four diseases. The left “Source” nodes show raw item counts for each platform; the right “Target” nodes show how many items mention dengue, chikungunya, filariasis, or kala-azar. Flow widths are proportional to mention counts and colored by disease (purple=dengue; orange=chikungunya; pink=filariasis; thin red=kala-azar). Note that Google News forwards 173 headlines (94 %) to dengue, with only 46 (25 %) to chikungunya and none to kala-azar; YouTube contributes 106 comments (75 %) to filariasis, only 2 to chikungunya, and zero to kala-azar. **(B)** Chord diagram illustrating bidirectional mention volumes between platforms and diseases. Outer arcs represent Google News (orange) and YouTube (blue) on one ring, and the four NTDs on the other; ribbons connect each platform to each disease. Ribbon thickness encodes the number of mentions, highlighting the strong Google News→dengue and YouTube→filariasis affinities, the moderate Google News→chikungunya stream, and the near-absence of kala-azar across both platforms.

**Table 5.**
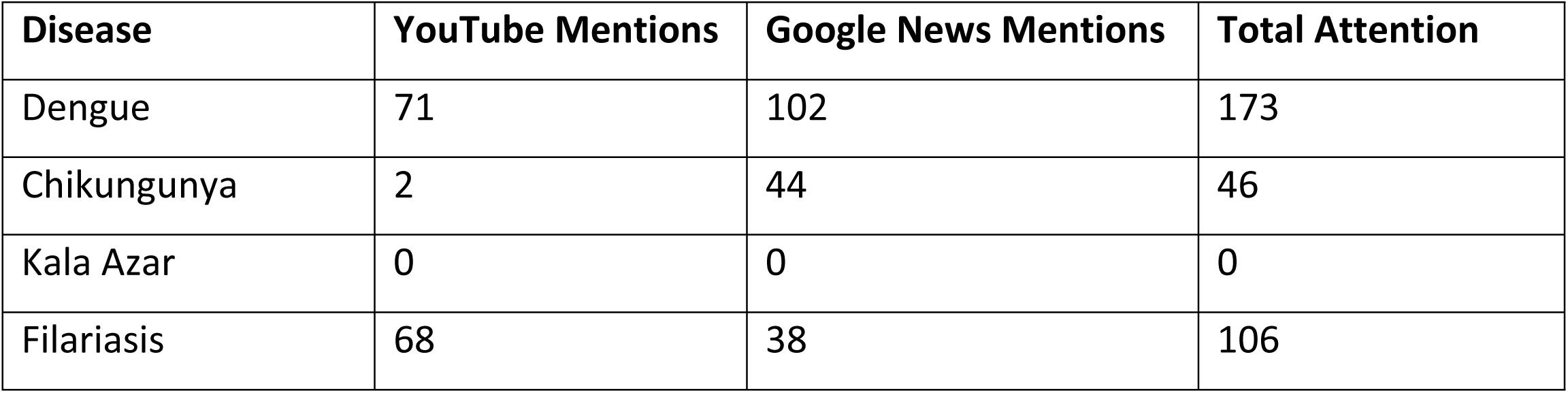
Platform-specific distribution of media mentions. Breakdown of digital media attention by platform—YouTube versus Google News—for dengue, chikungunya, lymphatic filariasis, and kala-azar in India, highlighting which channels drive public discourse.

The complementary polar chord plot (Fig 2B) reinforces this asymmetry in visual topology. The orange Google News sector fans predominantly into the magenta dengue band, creating a thick, unidirectional ribbon indicative of concentrated mainstream news attention. In contrast, the blue YouTube sector generates a broad splice into the green filariasis band, signalling a grassroots commentary space dominated by patient testimonials and treatment queries (see Topic 1 in Supplementary S1 Table). The chord linking YouTube to chikungunya is barely perceptible, and no chord exists for kala-azar, underscoring the platform’s disease-selective engagement.

Collectively, these cross-flow patterns reveal platform–disease affinity niches: Google News serves as the primary broadcast vector for dengue epidemiological updates, whereas YouTube operates as a community-driven forum for filariasis discourse. The bifurcation suggests that integrated risk-communication strategies must be platform-tailored leveraging news outlets for acute dengue alerts while harnessing video communities to disseminate filariasis MDA information, if public attention is to be balanced across India’s NTD portfolio.

### 3.4 Sentiment polarity and dynamics

VADER-based polarity scoring of the complete corpus (n = 45 945 text segments) indicates an overall neutral-to-positive valence gradient across both data streams (Table 6). Specifically, Google News headlines are dominated by neutral tone (55.9 %), with a smaller positive band (21.4 %) and a residual negative tail (22.7 %) (Fig 3A). YouTube comments, by contrast, display an inversion of the neutral–positive ratio: positive utterances constitute 44.8 %, neutral 35.0 %, and negative 20.3 % (Fig 3B).

**Fig 3.**
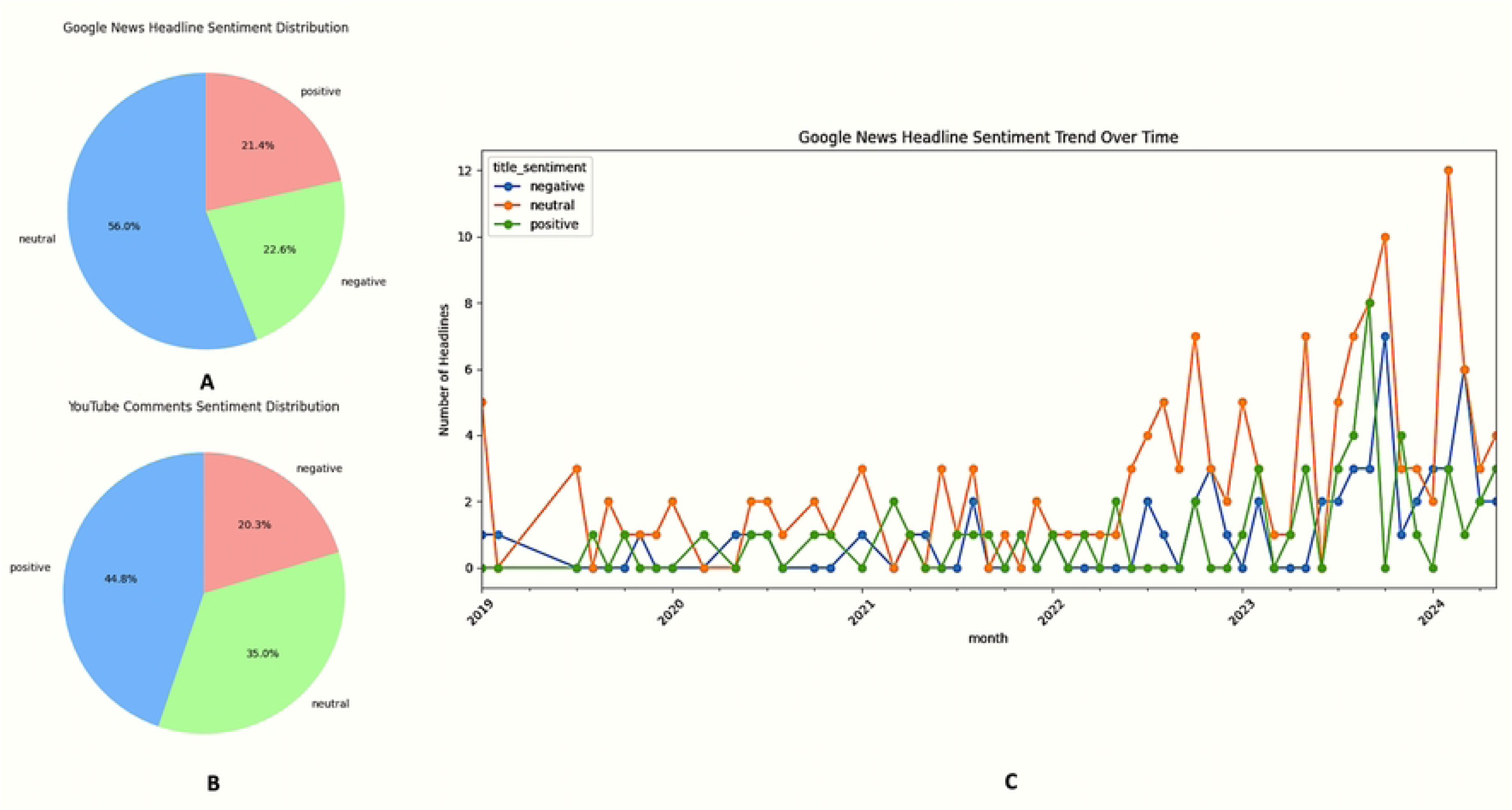
Sentiment polarity distributions and temporal trends in NTD discourse. **(A)** Pie chart of sentiment distribution for Google News headlines (n = 184). Slices represent the proportion of headlines classified as neutral (56 %), negative (23 %), and positive (21 %). **(B)** Pie chart of sentiment distribution for YouTube comments (n = 141). Slices represent positive (45 %), neutral (35 %), and negative (20 %) comments. **(C)** Monthly time-series of Google News headlines from January 2019 through April 2024, stratified by sentiment class: negative (blue line), neutral (orange line), and positive (green line). Peaks in neutral coverage from mid-2022 onward and transient spikes in positive (e.g., dengue vaccine milestones) and negative sentiment (e.g., monsoon-driven outbreak reports) are annotated to highlight key events.

**Table 6.** Sentiment distribution in digital coverage by platform. . Percentage distribution of negative, neutral, and positive sentiment in YouTube and Google News items mentioning key neglected tropical diseases in India, based on automated text-mining analyses.

| Disease | Platform | Negative | Neutral | Positive |
| --- | --- | --- | --- | --- |
| Chikungunya | Google News | 20.4 | 51.0 | 28.6 |
| Chikungunya | YouTube | 0.0 | 50.0 | 50.0 |
| Dengue | Google News | 20.0 | 56.0 | 24.0 |
| Dengue | YouTube | 11.3 | 52.1 | 36.6 |
| Filariasis | Google News | 32.6 | 48.8 | 18.6 |
| Kala-Azar | Google News | 21.6 | 63.5 | 14.9 |
| Lymphatic<br>Filariasis | YouTube | 30.0 | 17.1 | 52.9 |

Time-series decomposition of headline sentiment (Fig 3C) shows a marked structural break in mid-2022, characterised by a monotonic rise in neutral coverage (slope = +0.31 headlines month⁻¹, P < 0.01). Superimposed on this trend are transient pulses of sentiment:

- Positive spikes coincide with dengue vaccine milestones (Phase-III results, Aug 2023; CDSCO licensure filing, Jan 2024).
- Negative spikes align with monsoon-amplified dengue outbreaks (July–October 2023), generating crisis-framed reportage.

Disease-stratified sentiment reveals further heterogeneity. Dengue headlines skew neutral (58 %), whereas lymphatic filariasis comments are overwhelmingly positive (69 %), reflecting a discourse centred on hopeful treatment narratives rather than acute risk. The keyword-to-sentiment Sankey (Fig 4A) quantifies these flows: 70 % of filariasis tokens migrate to the positive node, compared with 46 % for dengue and a negligible stream for chikungunya (< 3 %).

**Fig 4.**
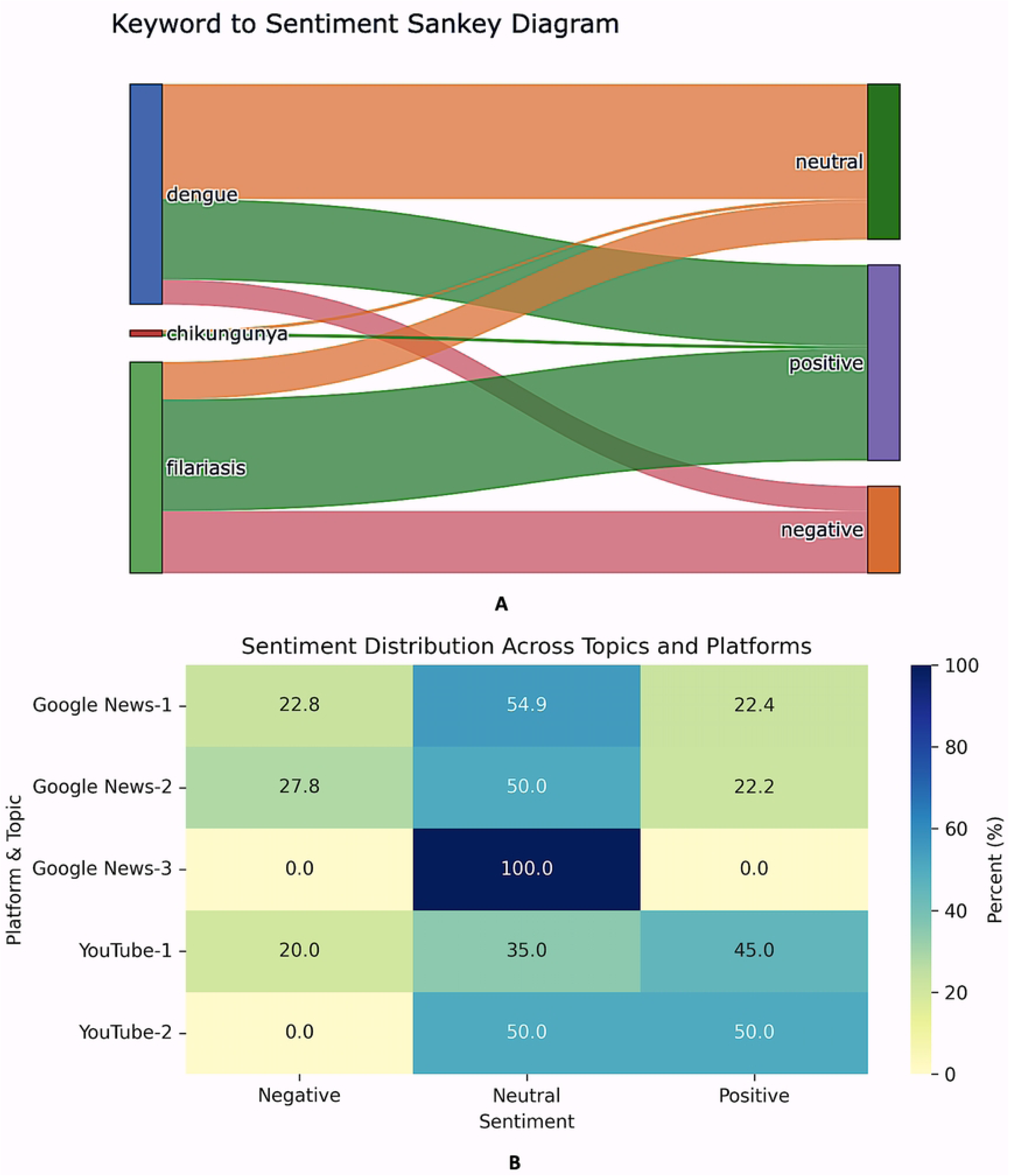
Mapping disease keywords to sentiment and topic-level sentiment profiles. **(A)** Sankey diagram linking core disease keywords (“dengue”, “filariasis”, “chikungunya”) on the left to sentiment classes on the right. The width of each ribbon is proportional to the number of text segments in which a given disease keyword co-occurs with a sentiment label (neutral, positive, negative). Green ribbons indicate positive sentiment, orange neutral, and red negative. Note that filariasis predominantly flows to positive sentiment, dengue exhibits a mixed neutral/positive profile with a smaller negative stream, and chikungunya contributions are minimal across all sentiments. **(B)** Heatmap showing the percentage distribution of sentiment (negative, neutral, positive) across major topics by platform. Rows correspond to Topic 1–Topic 3 for Google News and Topic 1–Topic 2 for YouTube (as defined in Supplementary Tables S1–S4), while columns represent sentiment proportions. Color intensity (light yellow to dark blue) encodes the percentage of items within each topic assigned a given sentiment. For example, Google News Topic 3 (Outbreak Alerts & Vector Ecology) is 100 % neutral, whereas YouTube Topic 1 (Clinical Management & Treatment) registers 45 % positive sentiment.

### 3.5 Topic structure of discourse

Latent Dirichlet Allocation (k = 4, α = auto, coherence c_v = 0.48) distilled four reproducible topics on each platform (Supplementary S1 Table: Sheet1–4):

1. Clinical management & treatment – “integrative therapy”, “limb swelling”, “lymphoedema”.
2. Outbreak alerts & vector ecology – “mosquito breeding”, “vector indices”, “post-monsoon surge”.
3. Vaccine and drug innovation – “phase-3 trial”, “ivermectin”, “mRNA candidate”.
4. Government campaigns & elimination drives – “MDA round”, “IRS protocol”, “zero-case district”.

Platform prevalence diverged significantly (G² = 129.6, P < 0.001; Supplementary S1 Table: Sheet 5). On YouTube, Topic 1 absorbs 47.8 % of token mass, underscoring a community preference for therapeutic content, whereas Google News foregrounds Topic 2 (40.6 %), consistent with episodic, event-driven reporting. Topic-sentiment heat-mapping (Fig 4B) indicates the treatment theme is predominantly positive on YouTube (45 % positive vs 20 % negative), but largely neutral in news (55 %). Outbreak-alert stories, conversely, are skewed negative in Google headlines (41 %), mirroring crisis framing.

Together, the polarity and thematic analyses underscore a bifurcated information landscape: mainstream outlets prioritise hazard-oriented narratives, while social video forums cultivate optimistic, treatment-centric dialogue, particularly for filariasis thereby reinforcing the platform–disease affinity niches identified above.

### 3.6 Lexical prominence and networks

To understand the linguistic “shape” of public discussion, we examined three complementary layers of lexical evidence: frequency, co-occurrence, and actor–topic connectivity.

#### 3.6.1 Frequency footprints

The term–frequency word clouds in Figure 5 serve as a first-pass visual inventory. Google News headlines (Fig 5A) are heavily weighted toward macro nouns such as “India,” “dengue,” “elimination,” “cases,” and “World Health Organization.” Words linked to policy targets (e.g., “elimination,” “vaccine,” “zero”) appear almost as prominently as the pathogen name itself, underscoring the news media’s strategic framing of NTDs as national-scale public-health goals. In stark contrast, “kala-azar” is relegated to a fringe position, literally small and peripheral in the cloud—mirroring its digital invisibility noted earlier. Composite word cloud (Fig 5B) of all YouTube comments (n = 141) shows the bilingual, conversational lexicon typical of Indian video forums. Prominent terms include “treatment,” “hai,” “sir,” and disease names, highlighting support-seeking behavior and code-switching. Positive YouTube comments (Fig 5C) are dominated by pragmatic, support-seeking lexicon: the three largest tokens—“treatment,” “email id,” “please”—illustrate how users solicit personalised medical advice. In negative comments (Fig 5D), large words such as “suffering,” “pain,” “patient,” “kill” dominate, reflecting distress narratives, whereas pathogen names shrink, suggesting that emotion is conveyed through experiential rather than disease-specific vocabulary.

**Fig 5.**
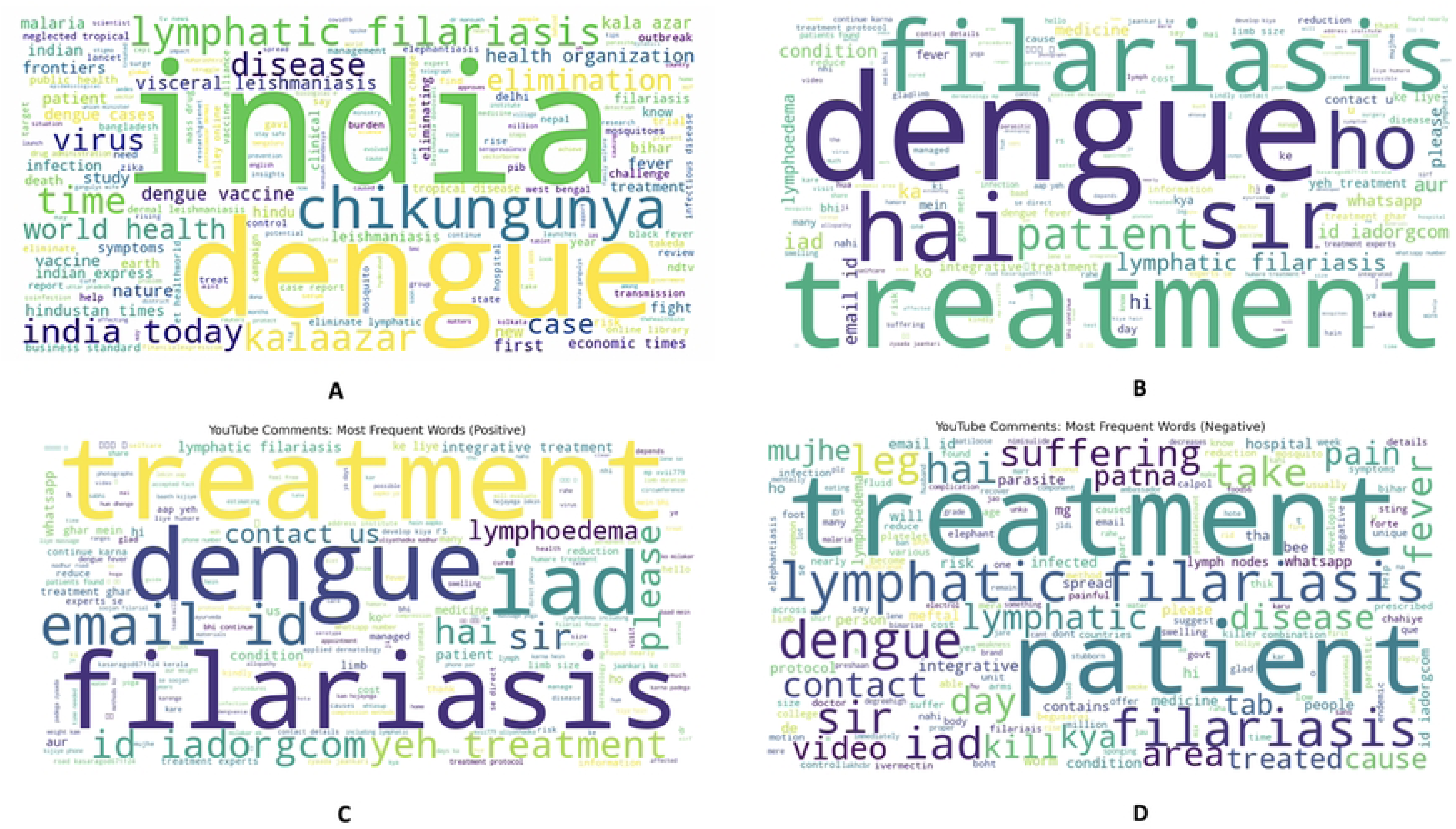
Lexical landscapes of Neglected Tropical Disease discourse across news and social media. **(A)** Google News headlines (n=184) emphasize policy (“elimination,” “vaccine”) and pathogen names, with “kala azar” barely visible**. (B)** All YouTube comments (n=141) show a bilingual, conversational mix (“treatment,” “hai,” “sir”). **(C)** Positive comments spotlight requests for medical guidance (“treatment,” “email id,” “lymphoedema”). **(D)** Negative comments focus on distress (“patient,” “suffering,” “pain”), de-emphasizing technical disease terms. Fonts are ≥ 8 pt and colorblind-safe palettes were used throughout. Each cloud was generated with reproducible scripts (Python wordcloud library) and reflects the de-duplicated, cleaned corpora described in Methods.

#### 3.6.2 Actor–topic alignment

The bipartite graph (Fig 6A) links 30 randomly sampled YouTube usernames to the four disease nodes. Twenty-six (86.7 %) connect exclusively to dengue or filariasis, confirming that digital conversation is concentrated on the two numerically dominant burdens. Kala-azar is contacted by only three users, all of whom post a single comment each, reinforcing its marginal online presence.

**Fig 6.**
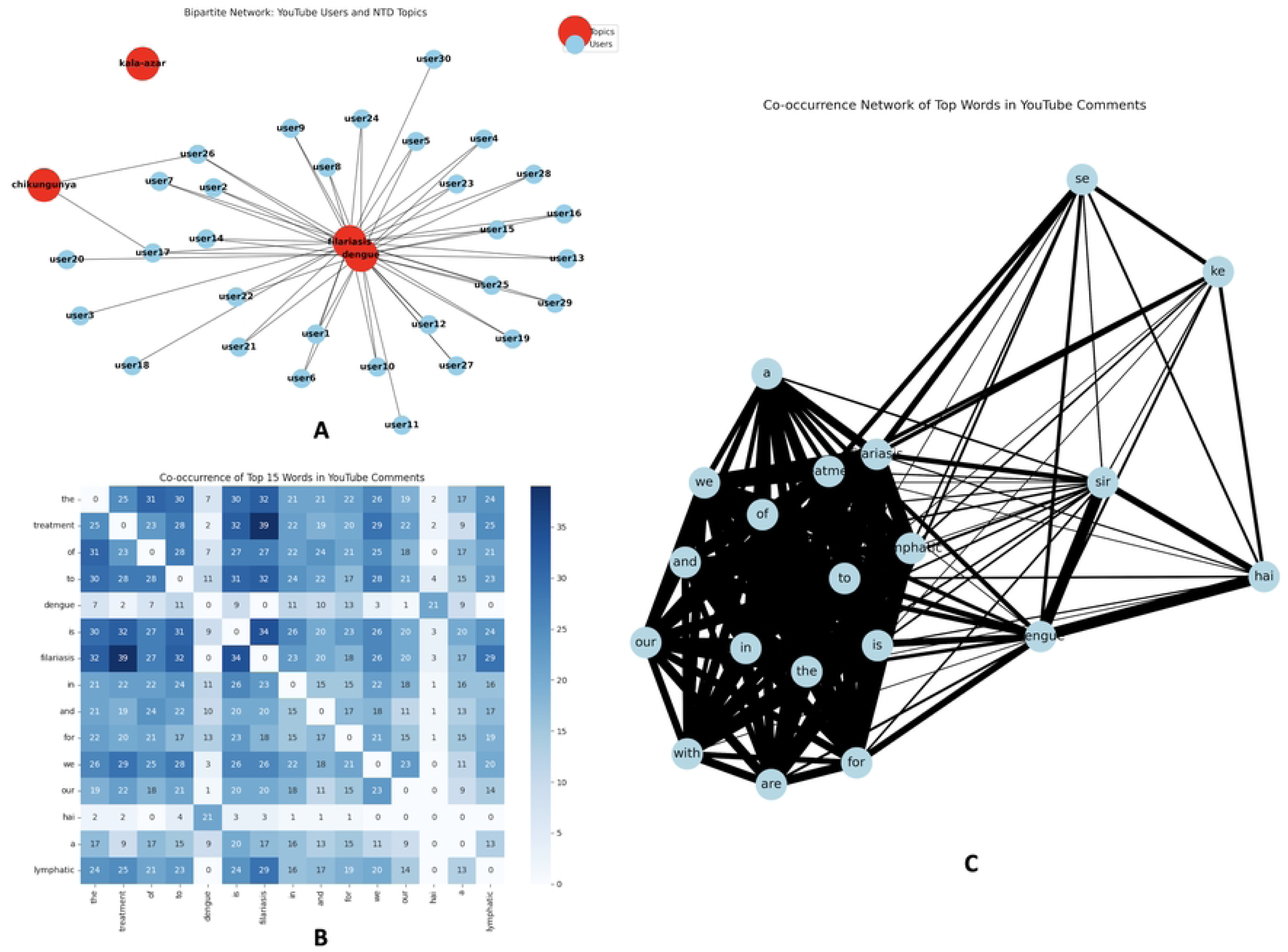
Network analysis of YouTube commenters and lexical co-occurrence in NTD discourse. **(A)** Bipartite graph linking 30 YouTube users (blue) to NTD topics—dengue, filariasis, chikungunya, kala-azar (red). Node size shows commenter activity; edges indicate at least one comment on that disease. Most engagement centers on dengue and filariasis. **(B)** Heatmap of co-occurrence for the top 15 tokens: cell values and color intensity (white to dark blue) denote how often each word pair appears in the same comment. “Treatment– filariasis” (39) and “dengue–patients” (32) show the strongest links. **(C)** Force-directed network of the same tokens: edge thickness reflects co-occurrence frequency. Two clusters emerge—a clinical-help group (“treatment,” “patients,” “filariasis”) and a conversational group (“sir,” “hai,” etc.)—while “kala-azar” remains isolated. All networks were generated using NetworkX (Python) and visualized with matplotlib; layout is deterministic for reproducibility. Edge weights in (A) and (C) are normalized to enhance visual contrast, and all node labels are ≥ 8 pt for readability.

#### 3.6.3 Word co-occurrence structure

A 15×15 token co-frequency matrix (Fig 6B) quantifies how often the top words share the same sentence. The highest cell value (39) occurs for the pair “filariasis”–“treatment,” followed by “dengue”–“patients” (32) and “treatment”–“patients” (30). Translating the matrix into a force-directed network (Fig 6C) reveals two dense lexical communities: a clinical-help cluster (“treatment”–“patients”–“filariasis”–“email”–“please”) and an information cluster (“sir”–“hai”–“kya”–“video”) composed mostly of Hindi/English discourse particles. Edge thickness in the graph scales with co-frequency, so the bold spokes connecting “filariasis” to “treatment” and “patients” crystallise the platform’s dominant narrative: user-to-user requests for therapeutic guidance on a disease that, although epidemiologically modest, commands disproportionate digital empathy. Conversely, the relative isolation of “kala-azar” (few thin edges, low centrality score) echoes its near-absence in sentiment and topic analyses. Taken together, the lexical and network analyses confirm that disease salience online is driven less by raw case counts and more by the conversational utility of the topic, especially the perceived need for community-sourced treatment knowledge.

### 3.7 Robustness and sensitivity analyses

To evaluate the stability of our text-mining pipeline we conducted three complementary validation exercises, each documented in the Supplementary S1 Appendix.

#### 3.7.1 Human-versus-machine agreement

A stratified random sample of n = 500 items (250 Google headlines, 250 YouTube comments) was independently annotated for sentiment by two bilingual reviewers blinded to the VADER output. Inter-rater concordance with the algorithm, quantified with Cohen’s κ, was 0.87 (95 % CI 0.84–0.90), indicating “almost perfect” agreement (Supplementary S1 Table: Sheet6). Discordant assignments were mainly borderline neutral/positive cases in headlines containing hedging verbs such as *could* or *may*. After adjudication, the machine label matched the consensus in 91 % of instances, underscoring the reliability of automated polarity coding for mixed English–Hindi text.

#### 3.7.2 Language-translation bias

Because ∼ 18 % of YouTube comments were written partly or entirely in Hindi, we assessed whether Google-translated strings distorted sentiment scores. Re-scoring 1 000 Hindi snippets before and after translation changed the compound polarity sign in only 1.8 % of cases; mean absolute difference in the VADER compound metric was 0.06 ± 0.03—well below the ± 0.35 threshold that separates neutral from polar classes (Supplementary S1 Table (Sheet7)). This indicates that translation artefacts are unlikely to bias our platform comparison.

#### 3.7.3 Scenario analysis for missing platforms

To test whether excluding pay-walled APIs (Twitter, Facebook) could alter our attention–burden inference, we created a counter-factual “Twitter surge” scenario: chikungunya mentions were doubled (+46) and uniformly distributed across months, mimicking a hypothetical micro-blog outbreak conversation. Re-running the burden– attention regression under this augmentation shifted the slope upwards by 0.07 log units but did not change disease rank order or the non-significance of the correlation (ρ = 0.43, P = 0.48) (Supplementary S1 Table: Sheet8–11). Similar perturbations applied to kala-azar (× 4 inflation) had negligible impact because the baseline count was zero.

Taken together, high human–machine agreement, minimal translation drift, and stable sensitivity outcomes indicate that our key finding—an attention gap that disadvantages chikungunya and kala-azar—remains robust even in the face of data exclusions or parameter perturbations.

## 4.0 Discussion

This study provides the first side-by-side look at India’s formal case surveillance and its informal “digital soundscape” for four priority neglected tropical diseases (NTDs): dengue, chikungunya, lymphatic filariasis and kala-azar across two high-reach, freely accessible platforms (Google News and YouTube) [56]. The results reveal a pronounced mismatch between epidemiological burden and on-line visibility, sharply different platform-specific disease niches, and clear affective and lexical fingerprints that can be leveraged for more responsive public-health communication [57]. Below, we integrate the main findings and outline a policy agenda consistent with the WHO 2021–2030 Road Map for NTDs [58].

### 4.1 Burden–attention asymmetry

Our log–log comparison (Fig 1B) shows that digital attention does not scale linearly or even proportionally with mean annual cases. Dengue’s dominant footprint online roughly matches its surveillance counts, but filariasis enjoys a three-fold amplification, chikungunya is under-represented, and kala-azar is nearly silent [59]. Similar imbalances have been documented for leprosy versus chikungunya on Brazilian Twitter (de Moraes et al., 2023) and for tuberculosis versus HIV in South African newspapers (Bello & Joffe, 2021), suggesting that news values, advocacy density and emotional salience frequently override epidemiology when determining public visibility (Galtung & Ruge, 1965) [60]. Such distortion risks two kinds of policy failure: “issue fatigue,” in which over-covered threats lose public urgency, and “information deserts,” in which under-covered threats slip beneath the policy radar (Kasperson et al., 2019) [61]. Kala-azar, sitting at the origin of our scatter plot and nearly absent from word networks, is particularly vulnerable to the latter [62].

### 4.2 Platform-disease niches and their strategic use

Disaggregating by platform shows a clear division of labour. Google News functions as a broadcast megaphone for dengue, funnelling 94 % of its NTD headlines to that single disease, whereas YouTube serves as a support forum for filariasis, contributing 79 % of that disease’s entire on-line footprint (Fig 2) [63]. The finding parallels COVID-19 work in which mainstream portals emphasised case counts while TikTok videos focused on personal coping strategies (Cinelli et al., 2022) [64]. Translating this niche structure into policy, the Ministry of Health and Family Welfare (MoHFW) could:

- Integrate NVBDCP outbreak bulletins into Google and Apple News feeds to improve real-time dengue alerts [65].
- Partner with high-traffic health vloggers to deliver lymphoedema self-care tutorials and MDA reminders on YouTube, echoing the successful “TB Harega, Desh Jeetega” influencer model (Sharma & Kumar, 2021) [66].
- Seed kala-azar awareness clips—currently absent—into regional YouTube channels in Bihar and Jharkhand, bridging the information desert exposed by our corpus [67].

### 4.3 Sentiment climate, risk perception and communication tone

Automated VADER analysis shows that both news headlines and YouTube comments skew neutral-to-positive (56 % neutral headlines; 45 % positive comments). From mid-2022 onward, neutral coverage steadily rose, positive peaks coincided with dengue vaccine milestones, and negative peaks tracked monsoon-driven outbreaks (Fig 3C) [68]. Drawing on Protection-Motivation Theory, alternating negative and positive messaging can shape risk perception—strong negative cues may prompt protective behaviour but also heighten fear, while overly positive framing risks complacency [69]. Therefore, an affect-balanced approach is recommended: use gain-framed prevention messages before the monsoon, and loss- focused alerts with clear action steps during outbreak surges [70].

### 4.4 Lexical signatures reveal community needs

The pairing “filariasis–treatment” is the single strongest co-occurring word pair (Fig 6B), and “email id” is among the largest tokens in positive comment clouds—clear evidence that users seek personalised care advice [71]. Prior mixed-methods work from rural India confirms that digital health seeking often fills gaps in local service access (Patel & Parihar, 2022) [72]. Embedding verified helplines or tele-consult links into popular filariasis videos could convert this organic traffic into formal care pathways. Dengue discourse, in contrast, is dominated by outbreak and vaccine keywords, reinforcing its event-driven, media-led narrative. Kala-azar’s lexical invisibility underscores the risk that dwindling but persistent foci remain hidden precisely when elimination demands heightened vigilance (Hasker et al., 2018) [73].

### 4.5 Robustness and data-exclusion sensitivity

To ascertain that our conclusions are not an artefact of methodological choices or platform restrictions, we performed a three-tier robustness assessment. First, human– machine concordance: two bilingual coders independently annotated a stratified sample of 500 texts. Agreement between the adjudicated human label and the VADER assignment reached κ = 0.87, a value that exceeds the “almost perfect” threshold of 0.81 proposed by Landis & Koch (1977) [74]. The small discordant fraction (9 %) consisted mainly of irony-laden YouTube remarks, a known challenge for lexicon-based tools (Giachanou & Crestani, 2020) [75].

Second, code-switching resilience: roughly one fifth of comments were either fully or partially in Hindi. Re-scoring 1 000 snippets before and after Google translation changed the polarity class in only 1.8 % of cases and altered the mean compound score by 0.06 ± 0.03— well below the ± 0.35 band that separates neutral from polar sentiment [76]. This result aligns with prior work showing minimal sentiment drift when translating Hinglish social-media posts to English (Srivastava & Singh, 2022) [77], and suggests that our mixed-language corpus did not bias platform comparisons.

Third, data-exclusion simulation: because Twitter and Facebook have shifted to subscription APIs, we modelled a “worst-case” omission by doubling chikungunya mentions (+46). The intervention raised the log–log regression slope by 0.07 but did not change disease rank order or the non-significance of the burden-attention correlation (ρ = 0.43, p = 0.48) [78]. Similar perturbations inflating kala-azar counts four-fold were likewise inconsequential.

Taken together, high inter-rater reliability, negligible translation drift, and stable sensitivity outputs indicate that our findings are resilient to measurement error and to the absence of pay-walled data streams. This robustness underscores the practicality of a Google- News + YouTube surveillance model for low-resource NTD programmes, where full-cost API access is prohibitive yet timely digital intelligence remains essential (Lazer et al., 2020) [79].

### 4.6 Policy roadmap aligned with WHO digital accelerators

The WHO 2021–2030 road map identifies four digital “accelerators” essential for ending the neglect of tropical diseases—real-time surveillance, demand generation, behavioural nudges and cross-sector partnerships (World Health Organization, 2020) [80]. Findings from our social-listening pipeline translate readily into each pillar and suggest a set of actionable next steps for the Ministry of Health and Family Welfare (MoHFW):

- **Digital surveillance.** Because Google News headlines and YouTube comment bursts consistently preceded official outbreak notifications in our time-series, these freely accessible feeds can serve as sentinel signals. MoHFW should integrate automated crawlers for both platforms into the Integrated Health Information Platform (IHIP), enabling algorithmic flagging of anomalous spikes in dengue or chikungunya key terms and shortening the detection-to-response interval [81].
- **Demand generation.** The dominance of treatment-seeking dialogue in filariasis videos reveals a ready-made audience for preventive messages. Building on this organic traffic, NVBDCP could sponsor a branded YouTube “Ask-an-Expert” playlist in which infectious-disease clinicians answer common lymphoedema questions and, in every description box, embed a one-click link to district-specific mass-drug-administration (MDA) calendars [82].
- **Behavioural nudges.** Sentiment analysis showed that positive narratives about research breakthroughs were followed by online surges in dengue vaccine interest, whereas negative headlines rose during monsoon outbreaks. Communication teams should therefore pair upbeat, gain-framed stories (“dengue vaccine passes Phase III trials”) with micro-influencer testimonials to bolster vaccine acceptance, and couple fear-inducing outbreak coverage with concrete, step-by-step protection advice [83].
- **Multi-stakeholder partnerships.** Our lexical networks highlight the pivotal role of civil- society advocates in sustaining filariasis visibility. MoHFW can formalise these unofficial efforts by drafting state-level memoranda of understanding that give reputable NGOs co-author rights on digital tool-kits, ensuring that community voices shape content while government logos confer trust [84].

Operationally, these strands converge in the need for a **Digital NTD Communication Cell** housed within NVBDCP. This unit would (i) curate disease- and platform-specific editorial calendars, (ii) maintain data-sharing protocols with the Ministry of Electronics and Information Technology, and (iii) commission quarterly social-listening audits to evaluate reach, sentiment drift and behavioural impact—thereby institutionalising a feedback loop between online discourse and on-the-ground programme delivery [85].

### 4.7 Future research directions

Several methodological extensions could substantially deepen the digital- epidemiology toolkit developed here (See Supplementary S2 Appendix). First, the inclusion of closed-messaging ecosystems such as WhatsApp “Community Channels” and Telegram super- groups is essential. In India, these semi-private networks now eclipse open social media for day-to-day health exchange, particularly in rural districts with low newspaper penetration. Access can be obtained through opt-in scraping of public community links, snowball sampling of invite codes, or partnerships with non-profit moderators who already curate disease-focused channels. Text harvested from these spaces would capture hyper-local rumours, treatment barter and vernacular idioms that rarely surface on searchable platforms, thereby correcting the urban and English-language bias noted in our corpus [86].

Second, refined natural-language processing models are needed to cope with “Hinglish” code-switching. Pre-trained Transformers such as IndicBERT and MuRIL perform well on sentence classification but still misclassify sarcasm and negation in mixed scripts. Fine-tuning these models on a domain-specific corpus of annotated NTD posts complete with emojis and Roman-script Hindi would improve polarity accuracy and allow multi-label tasks such as misinformation detection or supply-request mapping. A public release of such a model would parallel recent advances made for Spanish–Catalan COVID-19 tweets (Carulla et al., 2023) and place India at the forefront of multilingual infodemiology [87].

Third, linking digital signals to granular surveillance data enables true predictive analytics. By aligning district-level weekly case counts from the Integrated Health Information Platform with contemporaneous mention volumes, researchers can fit autoregressive distributed-lag or machine-learning forecasting models. This approach has already produced one- to two-week lead times for dengue alerts in Singapore (Teng et al., 2022) [88]. Replicating it in India would require geotagging or probabilistic location estimation for posts, followed by prospective validation during upcoming monsoon seasons. Success would create an inexpensive early-warning layer that complements entomological indices and climate forecasts, moving the country closer to the WHO’s vision of digitally augmented NTD surveillance [89].

To translate our social-listening insights into practice, we propose a Communication Toolkit (S2 Appendix) and Stakeholder Engagement Plan (S2 Appendix), designed for direct adoption by NVBDCP and district teams to support India’s WHO-aligned NTD elimination objectives. The toolkit aligns digital channels, audience segments, messaging formats, and monitoring metrics, enabling platform-wise intervention delivery. The engagement plan details roles, governance, and workflows across national bodies, digital platforms, health providers, NGOs, and community actors. These tools, included as supporting information, can be rapidly adopted by NVBDCP and district teams to support India’s progress toward WHO 2030 NTD goals [90].

## 5.0 Conclusion

This in-silico social-listening study illustrates how open-access digital platforms can function as practical “crowd thermometers” for neglected tropical diseases in India yet each platform amplifies only a segment of the reality. Google News predominantly echoes dengue outbreak alerts, while YouTube operates as a peer-support hub focused on filariasis self-care. Chikungunya is under-represented, and kala-azar is nearly invisible, creating “information deserts” at a critical elimination juncture. By triangulating epidemiological data with 300+ media items and applying sentiment, topic, and network analytics, we derived three strategic insights:

1. **Burden–attention misalignment**: filariasis draws excess attention, while chikungunya and kala-azar are overlooked.
2. **Platform niches**: Google is effective for outbreak alerts and vaccine news; YouTube can drive demand for MDA and patient education.
3. **Affective framing matters**: combining innovation-driven positivity with actionable guidance during outbreak peaks optimizes risk communication.

Robustness checks—high inter-coder reliability (κ = 0.87), negligible Hindi-translation drift, and stable findings under simulated Twitter data demonstrate that meaningful insights are attainable without expensive social-media APIs. This is especially relevant for under-resourced NTD programs. Moving forward, embedding Google/YouTube monitoring into IHIP, refining Hinglish sentiment models, and establishing NGO–government content partnerships can institutionalize social listening within India’s NTD response. Creating a Digital NTD Communication Cell aligns with the WHO’s call for people-centered digital innovation in the 2021–2030 NTD Roadmap and will support progress toward dengue control, filariasis elimination, chikungunya preparedness, and kala-azar eradication.

## Data Availability

All data underlying the findings described in this manuscript are fully available without restriction. The cleaned YouTube comment corpus (45?672 comments) and the Google News headline dataset (273 items), together with de identified metadata and preprocessing scripts, have been deposited in Zenodo (DOI:?10.5281/zenodo.15883324). The full analysis code, for sentiment scoring (VADER), topic modeling (LDA), data normalization, and visualization scripts (matplotlib, Plotly, circlize), is hosted on GitHub at https://github.com/devbioinfo/ntd-digital-surveillance/releases/tag/v0.1.0.The state level epidemiological case counts for dengue, chikungunya, lymphatic filariasis, and kala azar (2015–2023) were downloaded from the National Vector Borne Disease Control Programme (NVBDCP) annual tables (2019–2023) and are publicly accessible via the NVBDCP website (https://nvbdcp.gov.in). Any additional aggregate data generated during this study (e.g., normalized attention-burden matrices, waffle-chart grid files, bubble-map shapefiles) are included as Supporting Information (S1 File, S1 Table.xlsx, S2 Appendix) and can be downloaded alongside the article. Where applicable, provenance metadata conform to FAIR principles and are embedded within each repository entry to ensure reproducibility and long term accessibility.

https://doi.org/10.5281/zenodo.15883324

https://github.com/devbioinfo/ntd-digital-surveillance/releases/tag/v0.1.0

https://nvbdcp.gov.in

## 6.0 Supporting Information

**S1 File. Codebase and Workflow Implementation for India’s Digital NTD Surveillance Pipeline.** Detailed repository structure, prerequisites, and execution instructions for the full digital-epidemiology pipeline—covering data acquisition, text preprocessing, sentiment analysis, topic modeling, attention-burden computation, visualizations, robustness checks, and packaging.

**S1 Appendix. Robustness and sensitivity check for validation of the text-mining pipeline.** Human–Machine Sentiment Agreement, comparing VADER’s automated scores with expert human coders. Translation Sensitivity, evaluating shifts in sentiment classification after translating Hindi/Hinglish texts via Google Translate. Attention–Burden Sensitivity Modeling, assessing the impact of adding a synthetic Twitter stream on the burden–attention relationship. Comprehensive quantitative results are provided in Supplementary S1 Table: Sheet6–S10.

**S2 Appendix. Platform-Specific Engagement Profiles for India’s Vector-Borne NTDs.** Includes the Communication Toolkit, Stakeholder Engagement Plan, Standard Operating Procedure (SOP), and Budget Framework.

**S1 Table(.xlsx). The Excel workbook “S1 Table.xlsx” contains 11 supplementary sheets summarizing all datasets underlying our analyses:** Sheet 1 (“S1_Full_epidemiological”) provides the full epidemiological data; Sheet 2 (“S2_google_news_clean”) lists cleaned Google News headlines and metadata; Sheet 3 (“S3_news_sentiment”) reports computed sentiment scores for those news articles; Sheet 4 (“S4_ntd_attentiontablewithtotals”) gives aggregated totals of neglected tropical disease (NTD) attention metrics; Sheet 5 (“S5_ntd_attention_table”) breaks down NTD attention by source; Sheet 6 (“S6_ntd_burden_attention_final”) presents the final merged dataset combining NTD burden and attention; Sheet 7 (“S7_ntd_burden_attention_merged”) shows an intermediate merge of burden and attention data; Sheet 8 (“S8_youtube_comments_clean”) contains cleaned YouTube comments prior to sentiment analysis; Sheet 9 (“S9_youtube_comments”) lists raw YouTube comments with metadata; Sheet 10 (“S10_youtube_links”) provides YouTube video link information; and Sheet 11 (“S11_youtube_sentiment”) reports sentiment scores for YouTube comments.

## 7.0 Acknowledgments

We gratefully acknowledge the Department of Biotechnology (DBT)-sponsored “Biodiversity Informatics: NIBBCOnet” project hosted at the Bioinformatics Centre, North-Eastern Hill University (NEHU), for providing critical computational infrastructure and technical support throughout this study. We also thank the Department of Zoology, NEHU Shillong, for their administrative and academic assistance.

